# A pilot feasibility study on SARS-CoV-2 detection method based on nasopharyngeal lavage fluid

**DOI:** 10.1101/2021.06.14.21258619

**Authors:** Daniele Frezza, Cristoforo Fabbris, Leonardo Franz, Elisa Vian, Roberto Rigoli, Rosalba De Siati, Enzo Emanuelli, Bertinato Luigi, Paolo Boscolo-Rizzo, Giacomo Spinato

## Abstract

**Objective:** Nose and nasopharyngeal swab is the preferred and worldwide accepted method to detect the Severe Acute Respiratory Syndrome Coronavirus 2 (SARS-CoV-2) within the nose and nasopharynx. This method may be linked with possible difficulties, such as patient’s discomfort or complications. This paper shows a pilot study of SARS-CoV-2 detection with nasal and nasopharyngeal lavage fluids.

**Methods:** Nasal lavage fluid was collected from patients who were submitted to SARS-CoV-2 screening test, due to a preceding positive rapid antigen test. A control group was enrolled among healthcare professionals whose nasopharyngeal swab tested negative. Nasal lavages were performed using isotonic saline solution injected through a nasal fossa. Both lavage fluid and traditional nasopharyngeal swab were analyzed by real-time PCR and antigenic test.

**Results:** A total of 49 positive subjects were enrolled in the study. Results of the analysis on lavages and nasopharyngeal swabs were concordant for 48 cases, regardless of the antigenic and molecular test performed. RT-PCR resulted weakly positive at swab in one case and negative at lavage fluid. Among the control group (44 subjects) nasopharyngeal swab and lavage fluid analyses returned a negative result. Sensitivity of the molecular test based on nasal lavage fluid, compared to traditional nasal swab, was 97.7%, specificity was 100%, and accuracy was 98.9%, with high agreement (Cohen’s k, 0.978).

**Conclusion:** Nasal and nasopharyngeal lavages resulted to be highly reliable and well tolerated. A larger series is needed in order to confirm these results. This approach may potentially represent a valid alternative to the traditional swab method in selected cases.

## Introduction

The current pandemic related to severe acute respiratory syndrome coronavirus 2 (SARS-CoV-2) infection, which causes COVID-19 (COronaVIrus Disease 2019), requiring massive population testing, rises some issues regarding diagnostic accuracy and feasibility on large scale of the current sample collection techniques.

As it has been demonstrated, samples from nasopharynx have the highest chances to contain material suitable for SARS-CoV-2 detection [1]. Accordingly, nasopharyngeal swab is currently the first choice for SARS-CoV-2 infection diagnosis [2]. However, it requires a steep learning curve for healthcare providers and is operator dependent. The diagnostic accuracy of nasopharyngeal swabs depends indeed on their success in reaching the nasopharynx. Such procedure may be affected by both poor performing technique (also due to anatomical misconception) and morphological barriers in the route to nasopharynx, including septal deviation, leading to increased discomfort during procedure [3]. Patient’s compliance to nasopharyngeal swabs is critical, especially in planning massive population-based testing campaigns. Nasopharyngeal swab is not completely devoid of adverse events risk, like epistaxis, septal hematoma or abscess, retention of foreign bodies from broken swabs or even cerebrospinal fluid leak [4,5]. Possible mistakes among health-care professionals deriving from long-lasting sessions of swab procedures must be also considered, with relative consequences on reliability of results [6].

Therefore, alternatives to the traditional swab should be developed to collect nasopharyngeal samples. Collection of lavage fluid has long been employed as a technique to obtain biological samples from the lower respiratory tract.

The aim of this study was to preliminarily explore the feasibility and accuracy of nasal lavage as a method to collect samples from the nasopharynx for SARS-CoV-2 detection.

## Methods

The present study was approved by the ethics committee of Treviso and Belluno provinces (ethic vote: 871/CESC). Written informed consent was obtained by all participants enrolled in the study. We collected nasal lavage fluid from patients who were submitted to SARS-CoV-2 screening test who were positive on the first rapid antigen test performed on a nasopharyngeal swab and returned to the Treviso SARS-CoV-2 Screening Center to perform the confirmatory PCR-based test. A control group was enrolled among healthcare professionals whose nasopharyngeal swab tested negative.

Each patient underwent the standard nasopharyngeal swab for molecular analysis. Subsequently nasal lavages were performed using isotonic saline solution injected through a nasal fossa with device Lavonase Lab. The solution mixed with nasal and nasopharyngeal secretions, once evacuated from the other nostril, was collected directly in a sterile tube (details are shown in Figure 1). After nasopharyngeal swab and nasal lavage collection, each participant was asked to report their perception of procedure discomfort by a 100 mm Visual Analogic Scales (VAS), ranging from 0 (no discomfort) to 100 (maximal discomfort).

**Figure 1.**
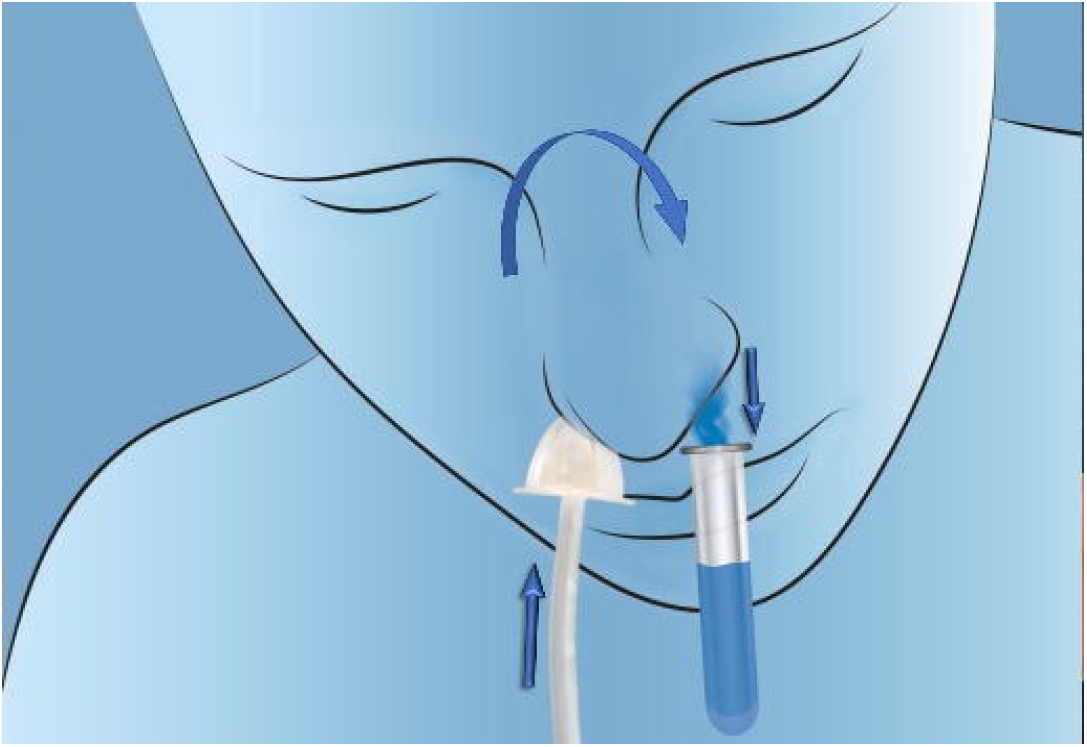
Scheme of the used nasal lavage. The head is bent down. The isotonic saline solution enters through a nasal fossa (upwards arrow), passes through the nasopharynx (curved arrow) and exits the other nasal fossa mixed with nasal and nasopharyngeal secretion, to be collected in the lab tube (downwards arrow) through a collection funnel.

**Figure 2.**
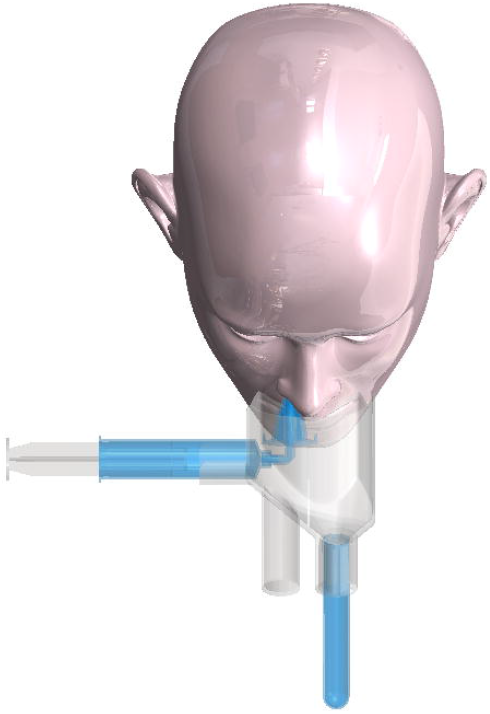
Example of the use of the device. A syringe is filled with isotonic sterile fluid (10 cc) and located on the right part of the device. The top of the syringe is linked with a cone-shaped rubber dispenser. The collection tube is inserted at the bottom in the left part. The patient bends down his head onto the device and introduces the top of the dispenser in his right nostril for a few millimeters. Then, he presses the plunger so that the fluid passes into the right nasal fossa, the nasopharynx, and the left nasal fossa. At the end, the fluid exits the left nasal fossa and collects into the test tube.

Both lavage fluid and traditional nasopharyngeal swab were analyzed by real-time PCR (*Allplex SARS-CoV-2 Assay, Seegene*) and antigenic test (*SARS-CoV-2 Rapid Antigen Test, Roche, F. Hoffmann-La Roche Ltd*) for SARS-CoV-2. For real-time -PCR analysis the considered detection limit to indicate negative samples was Ct⍰=⍰40.

Results of the traditional swab and the nasal lavage fluid analyses were compared calculating sensibility, specificity and accuracy and corresponding confidence intervals (CI). Agreement was evaluated through Cohen’s kappa. Difference in VAS discomfort between traditional swab and nasal lavage was evaluated through Mann-Whitney test.

## Results

All results are reported in Supplementary Table 1. The case group was composed of 49 consecutive subjects (mean [range] age, 49.0 [18-80] years; 30 [61.2%] male). In all subjects except one, the results of the analysis on lavage fluid and traditional nasopharyngeal swab were concordant, regardless of the antigenic and molecular test performed. Only in case n° 20 RT-PCR resulted weakly positive at swab (cycle threshold of E gene 37, cycle threshold of RdRP 38 and N genes 36) and negative at lavage fluid. A total of 44 subjects were enrolled in the control group (mean [range] age, 45.0 [26-61] years; 28 [63.6%] female). In all these cases, both the nasopharyngeal swab and the lavage fluid analyses returned a negative result. Therefore, the sensitivity of the molecular test based on nasal lavage fluid was 97.7% (95% CI: 88.2%-99.6%) while the specificity was 100% (95% CI: 92.7%-100%) resulting in an accuracy of 98.9% (95% CI: 94.2-99.8%) compared to the molecular test based in traditional nasal swab; high agreement was found (Cohen’s k, 0.978) (Table 1).

**Table 1.** Distribution of SARS-CoV-2 positive and negative cases among nasal swabs and lavages.

|  | Positive swabs | Negative swabs | Row total |
| --- | --- | --- | --- |
| Positive lavages | 43 | 0 | 43 |
| Negative lavages | 1 | 49 | 50 |
| Column total | 44 | 49 |  |

Overall, nasal lavage fluid collection was atraumatic and well tolerated with no adverse events. Collection of the lavage was performed twice in two patients because they did not sufficiently bend their head. Patients reported much lower discomfort with nasal lavage (median VAS: 0; interquartile range: 0-16) than with nasopharyngeal swab (median VAS: 68; interquartile range: 31-85; p<0.0001).

## Discussion

The diagnosis of SARS-CoV-2 infection based on RT-PCR from nasopharyngeal nasal lavage was observed to be both feasible and accurate with specificity and sensitivity being 97.7% and 100%, respectively, when compared with the gold standard PCR from nasopharyngeal swab.

The use of nasal lavage overcomes the critical issues risen by traditional nasopharyngeal swabs. First of all, such approach is easier to perform and more standardized, reducing the training time for the operators, or it can be performed by the patient himself with a reduction in the risk of contagion of COVID-19 for the operators and reduction of health costs. Moreover, issues related to collection, transport and storage could be greatly reduced [7].

Lavages might also allow to collect sufficient nasopharyngeal material even from anticoagulated patients or from those with severe septal deviation or with reduced compliance, like pediatric or non-cooperative subjects. Moreover, such approach avoids direct contact of a swab with nasal and nasopharyngeal mucosa. This is not only a compliance issue: nasal mucosa brushing, beside producing discomfort, damages the muco-ciliary clearance system, which may take several weeks to be repaired [8], leaving patients with less effective physical barriers against infectious agents.

The results in this clinical study preliminarily suggest an excellent tolerability and a minimal risk of operator-dependence. In selected cases, collection of nasal lavage fluids may be a potentially advantageous method to obtain samples from nasopharynx for SARS-CoV-2 detection and prevent COVID-19 spreading, compared to the traditional swab. However, this procedure should be more extensively investigated in larger cohort studies to evaluate the test-retest reliability, calculate limit of detection, and further assess patients’ compliance and safety from side effects.

## Conclusion

According to the present pilot clinical study, nasal and nasopharyngeal lavages resulted to be highly reliable and well tolerated, even if the research was conducted among a small case series. Further studies among a larger number of positive and negative subjects is needed in order to confirm these results. If these aspects will be confirmed, diagnosis based on nasal and nasopharyngeal lavage fluid may potentially represent a valid alternative to the traditional swab method.

## Data Availability

Data available on request

## Acknowledgements

The authors would like to acknowledge Jerry Polesel for his support in statistical analysis of the data.

## Supplementary Table legend

**Supplementary Table 1**. Series of patients who underwent nasal and nasopharyngeal lavage and nasopharyngeal swab analysis for SARS-CoV-2 detection. For RT-PCR analysis the considered detection limit to indicate negative samples was Ct⍰>⍰40.

