## Supplementary Table 1 for "A pilot feasibility study on SARS-CoV-2 detection method based on nasopharyngeal lavage fluid"

**Supplementary Table 1.** Series of subjects who underwent nasal lavage fluid and nasopharyngeal swab analysis for SarsCov2 detection. For RT-PCR analysis the considered detection limit to indicate negative samples was Ct > 40.

| Subject | Age | Gender | Antigenic analysis results | | Molecular (RT-PCR) analysis results | |
| --- | --- | --- | --- | --- | --- | --- |
|  |  |  | **Nasopharyngeal swab** | **Lavage fluid** | **Nasopharyngeal swab** | **Lavage fluid with** |
| 1 | 70’s | M | POSITIVE | POSITIVE | NEGATIVE | NEGATIVE |
| 2 | 50’s | M | POSITIVE | POSITIVE | POSITIVE | POSITIVE |
| 3 | 10’s | M | POSITIVE | POSITIVE | NEGATIVE | NEGATIVE |
| 4 | 80’s | F | POSITIVE | POSITIVE | NEGATIVE | NEGATIVE |
| 5 | 50’s | F | POSITIVE | POSITIVE | POSITIVE | POSITIVE |
| 6 | 30’s | M | POSITIVE | POSITIVE | POSITIVE | POSITIVE |
| 7 | 70’s | F | POSITIVE | POSITIVE | POSITIVE | POSITIVE |
| 8 | 40’s | F | POSITIVE | POSITIVE | POSITIVE | POSITIVE |
| 9 | 40’s | F | POSITIVE | POSITIVE | POSITIVE | POSITIVE |
| 10 | 50’s | M | POSITIVE | POSITIVE | POSITIVE | POSITIVE |
| 11 | 10’s | M | POSITIVE | POSITIVE | POSITIVE | POSITIVE |
| 12 | 70’s | M | POSITIVE | POSITIVE | NEGATIVE | NEGATIVE |
| 13 | 10’s | M | POSITIVE | POSITIVE | POSITIVE | POSITIVE |
| 14 | 60’s | F | POSITIVE | POSITIVE | POSITIVE | POSITIVE |
| 15 | 60’s | M | POSITIVE | POSITIVE | POSITIVE | POSITIVE |
| 16 | 60’s | M | POSITIVE | POSITIVE | POSITIVE | POSITIVE |
| 17 | 20’s | F | POSITIVE | POSITIVE | POSITIVE | POSITIVE |
| 18 | 20’s | M | POSITIVE | POSITIVE | POSITIVE | POSITIVE |
| 19 | 40’s | M | POSITIVE | POSITIVE | POSITIVE | POSITIVE |
| 20 | 70’s | M | POSITIVE | POSITIVE | **POSITIVE** | **NEGATIVE** |
| 21 | 20’s | M | POSITIVE | POSITIVE | POSITIVE | POSITIVE |
| 22 | 20’s | M | POSITIVE | POSITIVE | POSITIVE | POSITIVE |
| 23 | 50’s | M | POSITIVE | POSITIVE | NEGATIVE | NEGATIVE |
| 24 | 50’s | F | POSITIVE | POSITIVE | POSITIVE | POSITIVE |
| 25 | 10’s | M | POSITIVE | POSITIVE | POSITIVE | POSITIVE |
| 26 | 30’s | F | POSITIVE | POSITIVE | POSITIVE | POSITIVE |
| 27 | 40’s | F | POSITIVE | POSITIVE | POSITIVE | POSITIVE |
| 28 | 40’s | F | POSITIVE | POSITIVE | POSITIVE | POSITIVE |
| 29 | 30’s | F | POSITIVE | POSITIVE | POSITIVE | POSITIVE |
| 30 | 70’s | M | POSITIVE | POSITIVE | POSITIVE | POSITIVE |
| 31 | 50’s | F | POSITIVE | POSITIVE | POSITIVE | POSITIVE |
| 32 | 60’s | F | POSITIVE | POSITIVE | POSITIVE | POSITIVE |
| 33 | 30’s | M | POSITIVE | POSITIVE | POSITIVE | POSITIVE |
| 34 | 70’s | M | POSITIVE | POSITIVE | POSITIVE | POSITIVE |
| 35 | 60’s | M | POSITIVE | POSITIVE | POSITIVE | POSITIVE |
| 36 | 40’s | F | POSITIVE | POSITIVE | POSITIVE | POSITIVE |
| 37 | 60’s | F | POSITIVE | POSITIVE | POSITIVE | POSITIVE |
| 38 | 50’s | F | POSITIVE | POSITIVE | POSITIVE | POSITIVE |
| 39 | 60’s | M | POSITIVE | POSITIVE | POSITIVE | POSITIVE |
| 40 | 20’s | M | POSITIVE | POSITIVE | POSITIVE | POSITIVE |
| 41 | 40’s | M | POSITIVE | POSITIVE | POSITIVE | POSITIVE |
| 42 | 40’s | M | POSITIVE | POSITIVE | POSITIVE | POSITIVE |
| 43 | 40’s | F | POSITIVE | POSITIVE | POSITIVE | POSITIVE |
| 44 | 50’s | M | POSITIVE | POSITIVE | POSITIVE | POSITIVE |
| 45 | 50’s | M | POSITIVE | POSITIVE | POSITIVE | POSITIVE |
| 46 | 40’s | M | POSITIVE | POSITIVE | POSITIVE | POSITIVE |
| 47 | - | M | POSITIVE | POSITIVE | POSITIVE | POSITIVE |
| 48 | 40’s | F | POSITIVE | POSITIVE | POSITIVE | POSITIVE |
| 49 | 30’s | M | POSITIVE | POSITIVE | POSITIVE | POSITIVE |
| 50 | 30’s | F | - | - | NEGATIVE | NEGATIVE |
| 51 | 40’s | M | - | - | NEGATIVE | NEGATIVE |
| 52 | 30’s | M | - | - | NEGATIVE | NEGATIVE |
| 53 | 40’s | M | - | - | NEGATIVE | NEGATIVE |
| 54 | 30’s | F | - | - | NEGATIVE | NEGATIVE |
| 55 | 50’s | F | - | - | NEGATIVE | NEGATIVE |
| 56 | 50’s | F | - | - | NEGATIVE | NEGATIVE |
| 57 | 50’s | F | - | - | NEGATIVE | NEGATIVE |
| 58 | 20’s | F | - | - | NEGATIVE | NEGATIVE |
| 59 | 40’s | F | - | - | NEGATIVE | NEGATIVE |
| 60 | 30’s | F | - | - | NEGATIVE | NEGATIVE |
| 61 | 40’s | M | - | - | NEGATIVE | NEGATIVE |
| 62 | 50’s | F | - | - | NEGATIVE | NEGATIVE |
| 63 | 40’s | F | - | - | NEGATIVE | NEGATIVE |
| 64 | 50’s | F | - | - | NEGATIVE | NEGATIVE |
| 65 | 50’s | F | - | - | NEGATIVE | NEGATIVE |
| 66 | 50’s | F | - | - | NEGATIVE | NEGATIVE |
| 67 | 30’s | M | - | - | NEGATIVE | NEGATIVE |
| 68 | 20’s | M | - | - | NEGATIVE | NEGATIVE |
| 69 | 20’s | M | - | - | NEGATIVE | NEGATIVE |
| 70 | 20’s | F | - | - | NEGATIVE | NEGATIVE |
| 71 | 30’s | F | - | - | NEGATIVE | NEGATIVE |
| 72 | 50’s | F | - | - | NEGATIVE | NEGATIVE |
| 73 | 50’s | F | - | - | NEGATIVE | NEGATIVE |
| 74 | 30’s | M | - | - | NEGATIVE | NEGATIVE |
| 75 | 30’s | M | - | - | NEGATIVE | NEGATIVE |
| 76 | 30’s | F | - | - | NEGATIVE | NEGATIVE |
| 77 | 30’s | F | - | - | NEGATIVE | NEGATIVE |
| 78 | 40’s | M | - | - | NEGATIVE | NEGATIVE |
| 79 | 20’s | M | - | - | NEGATIVE | NEGATIVE |
| 80 | 40’s | F | - | - | NEGATIVE | NEGATIVE |
| 81 | 50’s | M | - | - | NEGATIVE | NEGATIVE |
| 82 | 50’s | F | - | - | NEGATIVE | NEGATIVE |
| 83 | 20’s | F | - | - | NEGATIVE | NEGATIVE |
| 84 | 40’s | F | - | - | NEGATIVE | NEGATIVE |
| 85 | 40’s | F | - | - | NEGATIVE | NEGATIVE |
| 86 | 60’s | M | - | - | NEGATIVE | NEGATIVE |
| 87 | 20’s | M | - | - | NEGATIVE | NEGATIVE |
| 88 | 50’s | F | - | - | NEGATIVE | NEGATIVE |
| 89 | 20’s | F | - | - | NEGATIVE | NEGATIVE |
| 90 | 20’s | M | - | - | NEGATIVE | NEGATIVE |
| 91 | 30’s | M | - | - | NEGATIVE | NEGATIVE |
| 92 | 60’s | F | - | - | NEGATIVE | NEGATIVE |
| 93 | 20’s | F | - | - | NEGATIVE | NEGATIVE |
